# Impact Of Electronic Health Record Interface Design On Unsafe Prescribing Of Ciclosporin, Tacrolimus and Diltiazem: A Cohort Study In English NHS Primary Care

**DOI:** 10.1101/19011957

**Authors:** Brian MacKenna, Seb Bacon, Alex J Walker, Helen J Curtis, Richard Croker, Ben Goldacre

## Abstract

**Background:** In England, national safety guidance recommends that ciclosporin, tacrolimus and diltiazem are prescribed by brand name due to their narrow therapeutic windows and, in the case of tacrolimus, to reduce the chance of organ transplantation rejection. Various small studies have shown that changes to electronic health records (EHR) interface can affect prescribing choices.

**Objectives:** Our objectives were to assess variation by EHR system in breaches of safety guidance around prescribing of ciclosporin, tacrolimus and diltiazem; and to conduct user-interface research into the causes of such breaches.

**Methods:** We carried out a retrospective cohort study using prescribing data in English primary care. Participants were English general practices and their respective electronic health records. The main outcome measures were (1) variation in ratio of breaching / adherent prescribing all practices (2) description of observations of EHR usage.

**Results:** A total of 2,575,411 prescriptions were issued in 2018 for ciclosporin, tacrolimus and diltiazem (over 60mg); of these, 316,119 prescriptions breached NHS guidance (12.3%). Breaches were most common amongst users of the EMIS EHR (in 23.2% of ciclosporin & tacrolimus prescriptions, and 22.7% of diltiazem prescriptions); but breaches were observed in all EHRs.

**Conclusion:** Design choices in EHR strongly influence safe prescribing of ciclosporin, tacrolimus and diltiazem; and breaches are prevalent in general practices in England. We recommend that all EHR vendors review their systems to increase safe prescribing of these medicines in line with national guidance. Almost all clinical practice is now mediated through an EHR system: further quantitative research into the effect of EHR design on clinical practice is long overdue.

## Introduction

Over 1.1 billion prescriptions are issued through primary care in England each year, at a cost of £8.8 billion in 2018.[1] The overwhelming majority of these prescriptions are generated through an electronic health record system,[2] where the clinician selects a medicine from a “picking list”; specifies directions (e.g. “once daily”); and then signs the prescription either electronically or on a printed copy. All NHS practices use one of four EHRs made available through NHS Digital.[3] We have previously described a small design choice in an EHR medicines selection screen which costs the NHS approximately £9.5million in one year across a wide range of drugs.[4]

Tacrolimus and ciclosporin are used in organ transplantation and other conditions such as rheumatoid arthritis, psoriasis and severe atopic dermatitis. Both medicines have a narrow therapeutic window: minor differences in blood levels have the potential to cause graft rejection reactions and switching between tacrolimus products has been associated with reports of toxicity and graft rejection. Therefore, the Medicines and Healthcare products Regulatory Agency (MHRA) recommend that tacrolimus and ciclosporin are prescribed and dispensed by brand name.[5,6] Similarly, diltiazem (a calcium channel blocker commonly used for conditions such as angina and mild to moderate hypertension) should be prescribed and dispensed by brand for preparations containing greater than 60mg, as different brands of modified-release formulations with over 60mg diltiazem may not have the same clinical effect.[7]

Our team delivers OpenPrescribing.net, a publicly funded and openly accessible data explorer for NHS primary care prescribing with 130,000 unique users in the past year. OpenPrescribing.net supports bespoke data queries alongside numerous predefined standard measures for safety, cost, and effectiveness, with data shared for every practice in England. Two of these standard measures assess compliance with the guidance to prescribe a branded preparation of ciclosporin, tacrolimus and diltiazem (over 60mg). Through our prior knowledge around EHR design, as clinicians and researchers engaged with software development, we suspected there may be a relationship between choice of EHR system and breaches of this safety guidance.

We therefore set out to: describe and map variation between practices and their parent clinical commissioning groups (CCGs) in breaches of safe prescribing guidance for ciclosporin, tacrolimus and diltiazem (over 60mg), and assess the impact of EHR choice on the proportion of safety breaches. Having identified one EHR system as being strongly associated with safety breaches, we conducted user-testing to explore the causes of this association. We aimed to produce a rapid report to support further investigations and any required software modifications to the EHRs, in light of the patient safety risk.

## Methods

### Study design

A retrospective cohort study in prescribing data from all English NHS general practices and CCGs, complemented with data on EHR deployment from NHS Digital; and user-testing of two commonly used EHRs by a senior pharmacist.

### Data Sources

We extracted data from the OpenPrescribing.net database. This imports openly accessible prescribing data from the large monthly files published by the NHS Business Services Authority which contain data on cost and items prescribed for each month, for every typical general practice and CCG in England since mid-2010.[8] The monthly prescribing datasets contain one row for each different medication and dose, in each prescribing organisation in NHS primary care in England, describing the number of items (i.e. prescriptions issued) and the total cost. These data are sourced from community pharmacy claims data and therefore contain all items that were dispensed. We extracted all available data for standard general practices, excluding other organisations such as prisons and hospitals, according to the NHS Digital dataset of practice characteristics and excluded practices that had not prescribed at least one item per measure. We excluded all other organisations such as prisons and hospitals. Data on EHRs and in which general practice they are deployed was extracted from a monthly file that is circulated by NHS Digital to interested parties and available on request.[8]

### Describe the prevalence of safety breaches by general practices in their implementation of brand prescribing of prescribing of ciclosporin, tacrolimus and diltiazem

We measured the number of safety breaches and created practice level deciles at each month for the measure (Table 1) of proportion of brand prescribing of ciclosporin, tacrolimus and diltiazem (over 60mg).

**Table 1.** OpenPrescribing Measures of generic ciclosporin, tacrolimus and diltiazem prescribing.

| **Measure** | **Definition** (full technical definitions are available on GitHub[9]) |
| --- | --- |
| Ciclosporin and Tacrolimus | Total items of generic ciclosporin and tacrolimus preparations, as a proportion of total items of all ciclosporin and tacrolimus items. |
| Diltiazem | Total items of generic diltiazem modified-release preparations, as a proportion of total items of all diltiazem |
|  | modified-release items (over 60mg). |

### Describe variation in safety breaches by EHR

We measured the proportion of safety breaches for prescribing of ciclosporin, tacrolimus and diltiazem for each of the four principle EHRs.

### Influence of principle EHR on breaches of safety guidance

We conducted mixed effects logistic regression, to determine the effect size of the EHR used, and the extent to which CCG membership and other factors affected these estimates. The independent variable was each prescription as a binary choice between generic and branded, while the main fixed effect variable was EHR vendor, with CCG membership as a random effect. Other practice factors were selected *a priori* based on their reasonable availability, as well as clinical judgement. They were: % of patients over 65; % of patients under 18; % of patients with a long term health condition; dispensing practice status; single handed practice status; rural/urbanness of practice location; index of multiple deprivation.

Where data for predictor variables were missing, these practices were excluded from the regression.

### EHR System User-Interface Evaluation

One senior pharmacist issued prescriptions in the EMIS and SystmOne computer systems to a test patient and observed the prompts.

### Software and Reproducibility

Data management was performed using Python and Google BigQuery, with analysis carried out using Stata 13.1 and Python. Data, as well as all code for data management and analysis is archived online.[10]

### Patient and Public Involvement

Our website OpenPrescribing.net, is an openly accessible data explorer for all NHS England primary care prescribing data, which receives a large volume of user feedback from professionals, patients and the public. This feedback is used to refine and prioritise our informatics tools and research activities. Patients were not formally involved in developing this specific study design.

## Results

### Prevalence of safety breaches

In 2018, 316,119 (12.3%) of prescriptions for ciclosporin, tacrolimus and diltiazem (over 60mg) breached prescribing safety guidance from a total of 2,575,411 prescriptions for these items. Of practices who prescribed ciclosporin or tacrolimus at least once, 2241(41.2%) breached safety guidance, while 5777 practices (80.4%) breached safety guidance with regards to diltiazem. The the ratio of safety breaches as a proportion of all prescribing for these items for practices was a 12.3% (10th - 90th percentile range = 0-47%) for diltiazem and 12.2% (10th - 90th percentile range = 0-67%).

### Variation in safety breaches by EHR

Figure 1 shows the mean values for the (a) ciclosporin and tacrolimus and (b) diltiazem safety breaches as a proportion of all prescribing for the respective items since January 2016. The rate of breaches was consistently highest in EMIS practices and in 2018, 23.2% of all ciclosporin and tacrolimus prescriptions issued from the EMIS EHR breached safety prescribing safety guidance while 22.7% diltiazem (over 60mg) prescriptions breached safety guidance.

**Figure 1.**
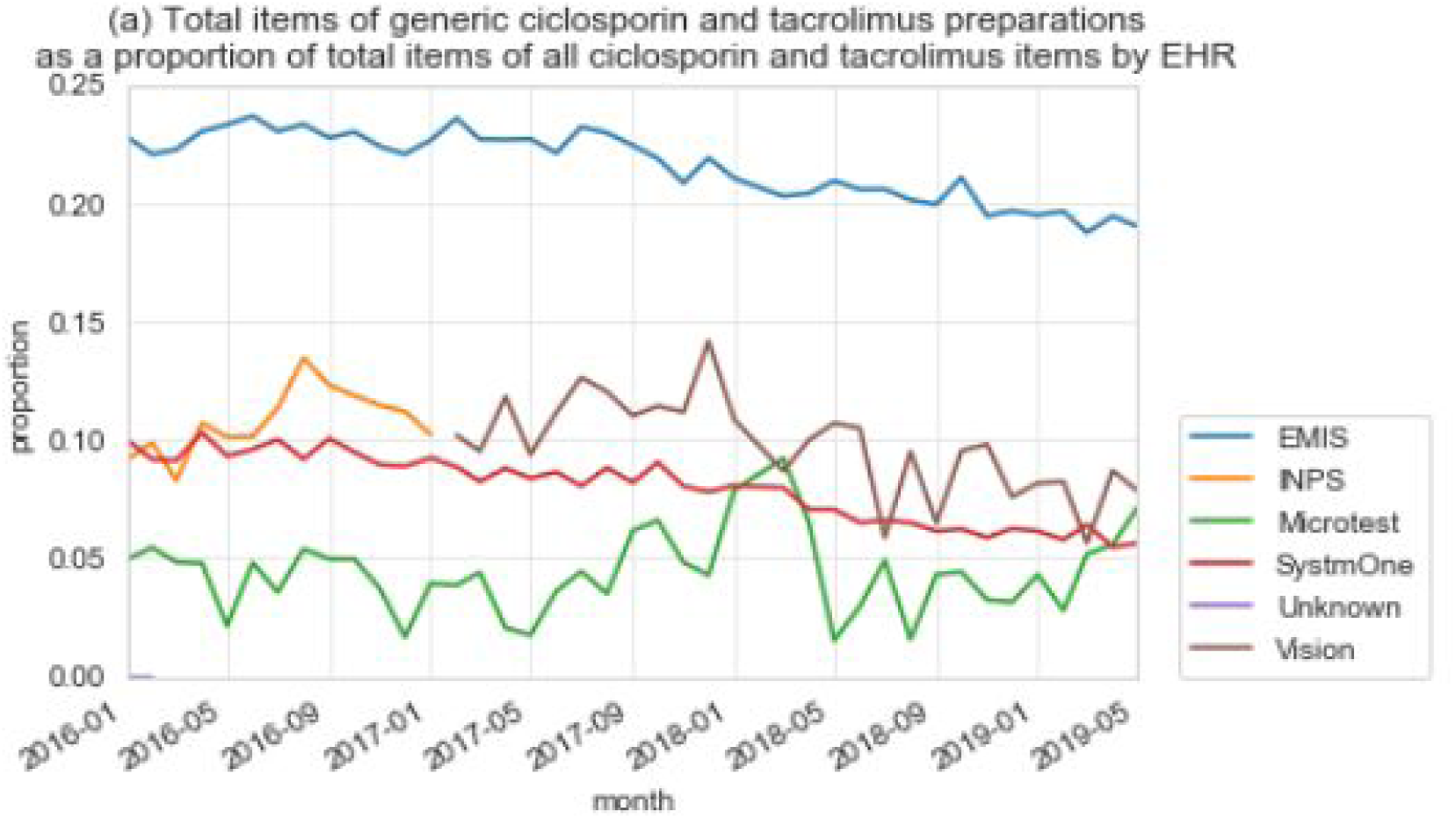
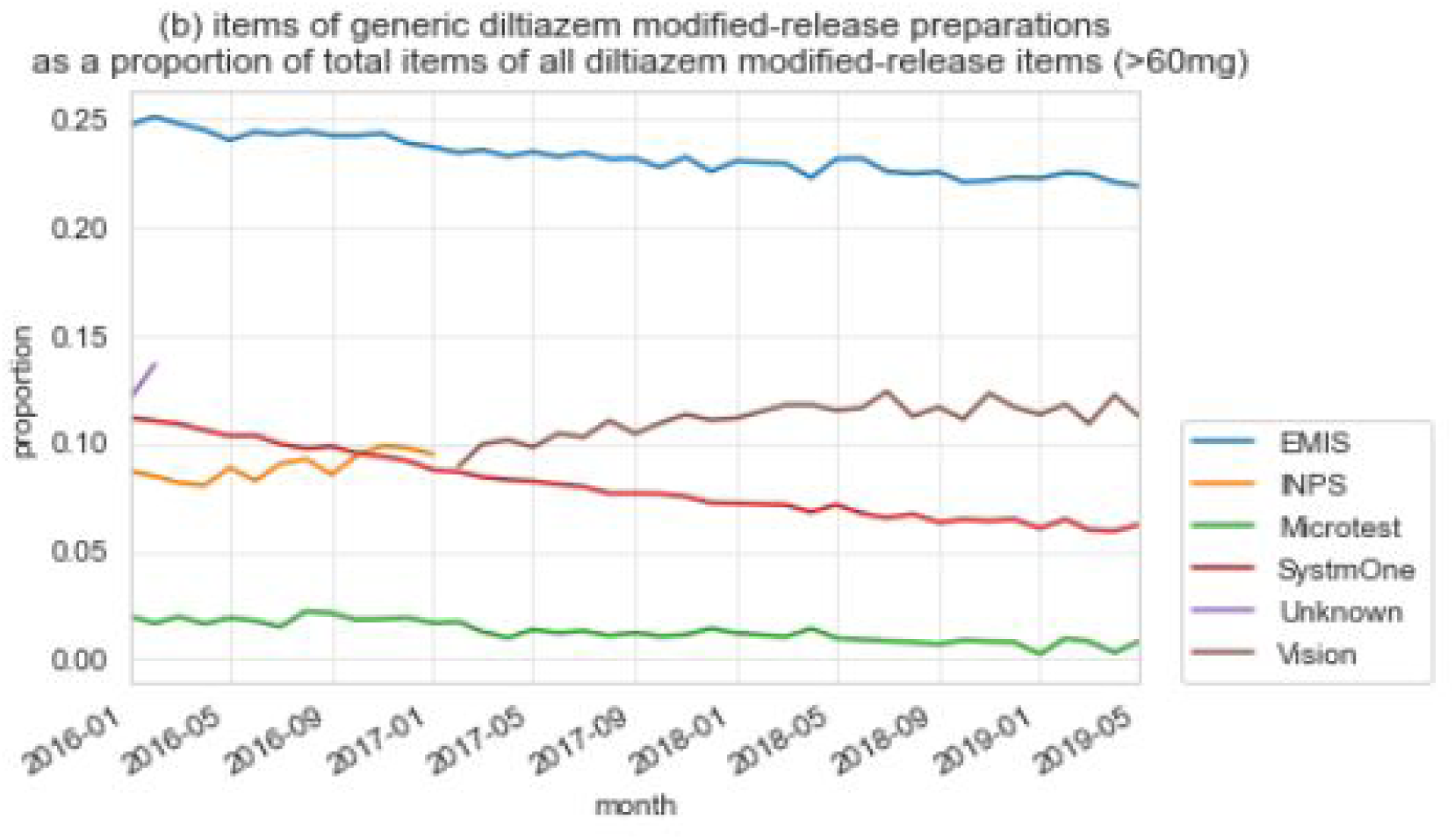
Mean values for ratio of safety breaches as a proportion of all prescribing by EHR.

The results of the logistic regression are shown in Table 2. When generating a diltiazem (over 60mg) prescription, practices using the EMIS EHR were around 4 times more likely to prescribe in breach of guidance with regards to diltiazem than practices using SystmOne, with a similar size of effect for ciclosporin and tacrolimus breaches. Practices using Microtest and Vision EHR were substantially less likely to breach safety guidance, although these practices make up a relatively small proportion of practices. Adjusting for CCG membership as a random effect had little effect on these estimates, although CCG membership was still responsible for a high proportion of the variation in propensity to prescribe generically (16.4% for diltiazem, and 8.4% for ciclosporin). Other factors had even less effect on the estimates, though some factors were weakly associated with the propensity to prescribe in breach of guidance (see <u>supplementary material</u>).

**Table 2.**
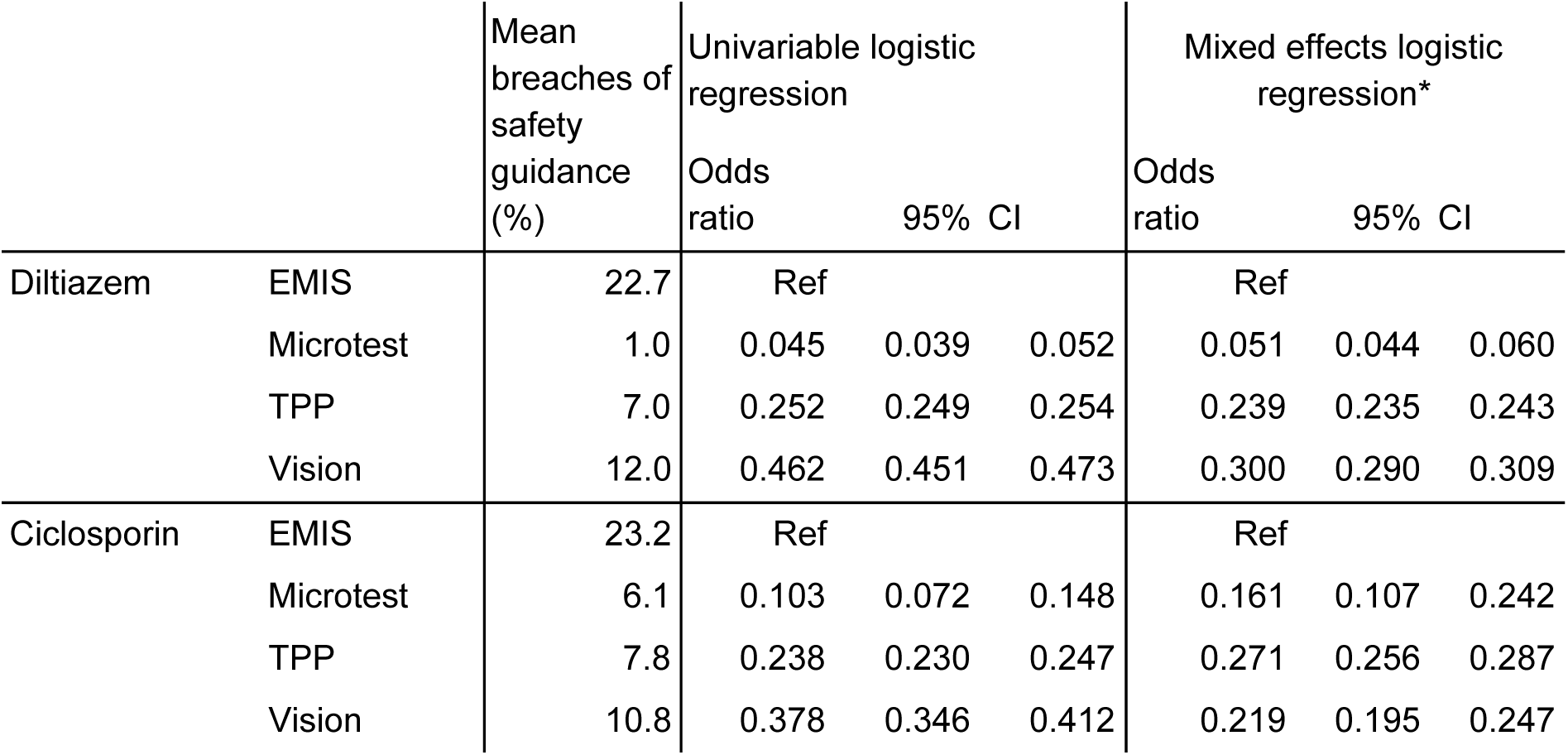
Ciclosporin and tacrolimus and diltiazem prescribing measures stratified by EHR along with odds ratios from a univariable and multivariable logistic regression model. *Adjusted for: CCG membership as a random effect; % of patients over 65; % of patients under 18; % of patients with a long term health condition; dispensing practice status; single handed practice status; rural/urbanness of location; index of multiple deprivation. (CI = Confidence Interval)

|  |  | Mean breaches of safety guidance (%) | Univariable logistic regression |  |  | Mixed effects logistic regression* |  |  |
| --- | --- | --- | --- | --- | --- | --- | --- | --- |
|  |  |  | Odds ratio | 95% CI |  | Odds ratio | 95% CI |  |
| Diltiazem | EMIS | 22.7 | Ref |  |  | Ref |  |  |
|  | Microtest | 1.0 | 0.045 | 0.039 | 0.052 | 0.051 | 0.044 | 0.060 |
|  | TPP | 7.0 | 0.252 | 0.249 | 0.254 | 0.239 | 0.235 | 0.243 |
|  | Vision | 12.0 | 0.462 | 0.451 | 0.473 | 0.300 | 0.290 | 0.309 |
| Ciclosporin | EMIS | 23.2 | Ref |  |  | Ref |  |  |
|  | Microtest | 6.1 | 0.103 | 0.072 | 0.148 | 0.161 | 0.107 | 0.242 |
|  | TPP | 7.8 | 0.238 | 0.230 | 0.247 | 0.271 | 0.256 | 0.287 |
|  | Vision | 10.8 | 0.378 | 0.346 | 0.412 | 0.219 | 0.195 | 0.247 |

### EHR System User-Interface Evaluation

#### EMIS

On reviewing the medicines selection screen it was observed that when searching for a medicine by its brand name, EMIS always presents the generic version of the medicine as priority in the medicines picking list: this represents a breach of safety guidance for ciclosporin, tacrolimus and diltiazem (over 60mg). Warnings are presented to prescribers to prescribe by brand when a generic version of these medicines is selected; however these warnings appear alongside multiple other warnings such as interactions with other medications. These warnings take the form of a “pop up” box which can be easily overridden and the generic version of all three medicines issued.

#### SystmOne

When the search terms “ciclosporin”, “tacrolimus” or “diltiazem” were inputted, no results are returned, encouraging a user to search by brand names in line with safety guidance. However in another part of the user interface screen there is a separate tick-box, “non-prescribable”. When ticked by a user, input of search terms “ciclosporin”, “tacrolimus” or “diltiazem” does return results allowing selection of the medicine generically in breach of safety guidance. A single, standalone “pop up” is presented if the user proceeds, warning of the importance of brand prescribing. This is in contrast to EMIS were it is presented alongside other clinical warnings.

## Discussion

### Summary

A total of 2,575,411 prescriptions were issued in 2018 for ciclosporin, tacrolimus and diltiazem (over 60mg); of these 316,119 prescriptions breached NHS safety guidance (12.3%). Breaches were most common amongst users of the EMIS EHR (numbers); but breaches were observed in all EHRs.

### Strengths and weaknesses

We included all typical practices in England, thus minimising the potential for obtaining a biased sample. We used prescribing data derived from pharmacy claims data used to calculate the transfer of funds from CCGs to dispensing pharmacies: all parties are motivated to ensure the accuracy of this data. We excluded a small number of settings such as walk-in centres, which typically do not issue repeat prescriptions for medicines, and where no data on EHR usage is available. The data does not distinguish which prescriptions may have been written by hand without EHR generation; however as 74% of prescriptions are transmitted electronically [2] and many more generated electronically and printed on paper, we do not expect this to substantially affect our results. Our data do not include hospital medicines data, but we do not expect the same issue associated with EHR design to occur in hospitals, as medicines are procured differently and the use of electronic prescribing and EHRs in secondary care in England is limited.[11] Observation of EHR was limited to one 30 minute session on each EHR with limited scenarios in a single location. EHRs can be manually adapted and defaults set locally; compliance with safety guidance may therefore differ between individual practices even with the same EHR systems.

One caveat regarding clinical impact is that the available data reflects prescribing and not dispensing: in England, when medicines are prescribed generically, pharmacies are entitled to dispense a brand, and some pharmacists may try to ensure they dispense the same brand that the patient previously received; however pharmacists are likely to be blocked in this by lack of access to the patient record, and, commonly, by the absence of information on specific brand in the patient’s record.[12] In any case occasional remedial interventions do not affect the key observation that EHR strongly predicts safety breaches among prescribing clinicians.

### Findings in Context

We are aware of no prior work on the prevalence of safety breaches around use of ciclosporin, tacrolimus and diltiazem: however, incomplete implementation of this important national prescribing safety recommendation is consistent with extensive prior work showing incomplete or slow adoption for other national prescribing guidance.[13–15] To our knowledge we are the first group to use natural variation in prescribing behaviour between EHR systems to identify, explain, and address suboptimal prescribing. A 2017 systematic review [16] identified 34 relevant studies exploring the role of computerised systems in suboptimal prescribing. However, none used quantitative methods to compare different systems, instead relying on questionnaires to interrogate clinicians about their experiences of EHRs; qualitative research observing or interviewing clinicians; and descriptions of clinicians’ spontaneous reports on errors and safety issues in EHR systems.

Various small studies have aimed to evaluate the impact of a single specific new change to a computerised prescribing system, as a behavioural intervention to increase the probability of a desired choice being made by clinicians using the system.[17–21] Despite evidence for effectiveness from “pop-ups”, “alert-fatigue” has also been described, whereby large numbers of pop-ups can result in salient messages being ignored or disabled by prescribers.[22] One small mix-method study involving the EMIS EHR found that only three “pop-ups” from 117 alerts resulted in the GP checking (but not altering) the prescription.[23]

### Policy Implications and Interpretation

Breaches of safety guidance around diltiazem, ciclosporin and tacrolimus can expose patients to avoidable clinical risk. This is especially important in the case of organ transplantation where failure to adhere to the safety guidance assessed in our analysis has resulted in organ rejection [6]. The current World Health Organisation challenge on medication safety encourages countries to reduce medication errors by 50% by 2022: our finding represents a clear example of an error amenable to change.[24] We strongly recommend that picking lists are configured in line with best practice guidelines, and that compliance with this is audited by national organisations such as NHS Digital and NHSX. The US National Coordinator for Health Information Technology has made similar recommendations.[16,25,26] For the specific safety issue raised by our analysis, the mandated medicines data standard for the NHS (DM+D) already includes a field that can be used to mandate branded prescribing where this is required by safety guidance; indeed it is likely that this field triggers the pop-up warnings described above.

### Future Research

We are concerned by the relative absence of applied practical research around the EHR systems used by clinicians to store information, retrieve relevant information rapidly when assessing a patient, and implement specific clinical actions such as ordering a test or prescribing a treatment. Healthcare activity is increasingly computerised, and EHR software is likely to exert a very substantial influence on the way that modern medicine is practiced, in the same way that the rapid explosion in social media usage has changed the ways that people interact socially.[27] We can find no previous attempts to evaluate the impact of EHR design on clinical practice by analysing variation between organisations using different EHR systems. In our view questions of how best to represent, retrieve, and present knowledge about patients to clinicians - and the impact this can have on patient care - should be a key priority for funders and researchers in “digital health”. The NHS spends large sums of money on EHRs [28] and there is substantial room for collaborative improvement to make clinical care safer, cheaper, and more effective.

There is also a role for open standards in this work. We are aware that in most general practices, complementary decision support systems are deployed alongside the core HER system, to support medicines optimisation activity and other aspects of quality improvement. [29–31] However we have been repeatedly blocked from researching the impact of these “pop-ups” on routine prescribing as there is no national framework or data detailing which pop-ups are implemented in each setting, and we have been unable to access the data by private negotiation. NHS commissioners and leaders are also blocked from routinely monitoring which pop-ups are implemented across the NHS; and from defining or deploying pop-ups nationally. In our view this represents a failure to set standards and ensure the proportionate and secure data-sharing necessary to evaluate and improve patient safety.

## Summary

National guidance on safe prescribing for ciclosporin, tacrolimus and diltiazem is commonly breached; and the prevalence of safety breaches is strongly influenced by the brand of electronic health record system. We recommend that EHR vendors immediately review their systems to mandate safe prescribing of these medicines in line with national guidance; and that more data and funding is made available to support research into the impact of EHR design on clinical practice.

## Data Availability

Data are available in a public, open access repository on Github.

https://github.com/ebmdatalab/software_differences_paper

## Acknowledgements

We are grateful to Peter Inglesby and Dave Evans for their contribution to maintaining databases and the OpenPrescribing website. We are grateful to wider NHS colleagues for discussions which have informed our work on this topic.

## Ethical approval

This study uses open, publicly available data, and data publicly available from NHS Digital on request therefore no ethical approval was required.

## Data Availability

Data are available in a public, open access repository on Github.[10]

## Contributorship

All authors conceived the study and designed the methods. BMK SB AW analysed the data. BMK HC BG drafted the manuscript. All authors contributed to and approved the final manuscript. BG supervised the project and is guarantor.

## Abbreviations

CCG: Clinical Commissioning Group
EHR: Electronic Health Records
MHRA: Medicines and Healthcare products Regulatory Agency
NHS: National Health Service

